# Impact of GLP-1 Receptor Agonists on Chronic Low Back Pain in Patients with Obesity: A Prospective Pilot Cohort Study

**DOI:** 10.64898/2026.05.20.26353666

**Authors:** Braeden Benedict, Dyan White-Gilliam, Aryan Pradhan, Salim Yakdan, Ahmad Hammo, Lucas Budd, Faraz Arkam, Simon Y. Tang, Kenneth B. Schechtman, Abby L. Cheng, Susan Robinson Reeds, Burel R. Goodin, Jacob K. Greenberg

**Affiliations:** Department of Neurological Surgery, Washington University School of Medicine in St. Louis, St. Louis, Missouri, USA; Department of Anesthesiology, Washington University School of Medicine in St. Louis, St. Louis, Missouri, USA; Department of Orthopaedic Surgery, Washington University School of Medicine in St. Louis, St. Louis, Missouri, USA; Division of Biostatistics, Washington University School of Medicine in St. Louis, St. Louis, Missouri, USA; Department of Medicine, Washington University School of Medicine in St. Louis, St. Louis, Missouri, USA

**Author notes:** Corresponding author: Jacob K. Greenberg, Address: 660 S. Euclid Ave, Campus Box 8057, St. Louis, MO 63110.

**Keywords:** Chronic low back pain, back pain, obesity, GLP-1 receptor agonists, weight loss, inflammation

## Abstract

**Objective:** To evaluate whether glucagon-like peptide-1 receptor agonists (GLP-1 RAs) are associated with improvements in pain severity, disability, quality of life, and physical function in adults with obesity and chronic low back pain (cLBP), and to explore potential mechanisms.

**Design:** Prospective, single-arm cohort study.

**Subjects:** Thirty-five adults (median age 41 years; 86% women) with obesity (median BMI 39.9 kg/m²) and cLBP initiating GLP-1 RAs (tirzepatide, n=24; semaglutide, n=11).

**Methods:** Participants completed questionnaires at baseline, 3, 6, 9, and 12 months. The primary outcome was Brief Pain Inventory-Short Form (BPI-SF) pain severity. Secondary outcomes included body mass index (BMI), BPI-SF pain interference, Numerical Rating Scale (NRS) back pain, Oswestry Disability Index (ODI), and Short Form-12 (SF-12). At baseline and 6 months, a subset (n=24) underwent quantitative sensory testing, physical performance testing, and blood draws for inflammatory biomarkers (C-reactive protein, TNF-α, IL-6, IL-10), adipokines (leptin, adiponectin), and hemoglobin A1c.

**Results:** Over 12 months, BMI decreased by 12.5% (median 39.9 to 34.9 kg/m², 95% CI [-6.6, -4.2]). BPI-SF pain severity improved (median 4.8 to 2.0, 95% CI [-2.1, -0.8]), as did pain interference, ODI, NRS back pain, and SF-12 physical component scores. Hemoglobin A1c, leptin, and C-reactive protein decreased. Adiponectin increased and physical performance improved, but neither reached significance. Experimental pain sensitivity was unchanged.

**Conclusions:** GLP-1 RAs were associated with clinically meaningful improvements in pain, disability, and quality of life. These findings suggest GLP-1 RAs may be a promising nonsurgical therapy for cLBP; randomized controlled trials are needed to establish causality and mechanisms.

## Introduction

Affecting more than 600 million people, chronic low back pain (cLBP) is the single greatest cause of years lived with disability worldwide,^1^ is a major source of healthcare spending,^2^ and accounts for billions of dollars in lost worker productivity.^3^ Despite its profound prevalence and impact, effective treatments for nonspecific cLBP are lacking, with low quality evidence and weak effect sizes for most nonsurgical treatments.^4^

Obesity is a key modifiable risk factor for cLBP.^1,5^ High body mass index (BMI) is estimated to be responsible for 36 million prevalent cases of low back pain and more than $23 billion in healthcare expenditures globally.^6^ Individuals with obesity are at higher risk for structural manifestations of lumbar degeneration,^7^ such as disc herniation.^8^ While increased biomechanical loading is one potential etiology for this pain, obesity has also been linked to both osteoarthritis development and pain sensitivity.^9^ Furthermore, adipokines such as leptin have been associated with intervertebral disc degeneration,^10^ and pro-inflammatory biomarkers have been associated with nonspecific low back pain.^11^ Despite this evidence linking obesity to cLBP development, the extent to which obesity treatment improves low back pain symptoms is unclear. For example, limited evidence suggests that bariatric surgery may reduce back pain in patients with severe obesity.^12^ However, broader evidence is limited, partly due to the difficulty of achieving reliable weight loss.^13,14^

Glucagon-like peptide-1 receptor agonists (GLP-1 RAs), such as semaglutide, and the dual GLP-1/glucose-dependent insulinotropic polypeptide (GIP) agonist, tirzepatide, were initially approved for type 2 diabetes but have now revolutionized the medical management of overweight and obesity.^15,16^ A series of randomized controlled trials (RCTs) have established the efficacy of GLP-1 RAs in producing significant weight loss and improving other health outcomes, including cardiovascular disease and kidney disease.^17^ These medications have also been associated with anti-inflammatory effects, though the extent to which their observed benefits are related to direct anti-inflammatory effects, beyond their impact on weight and adiposity, remains poorly defined.^18^

Growing evidence suggests that GLP-1 RAs may have therapeutic benefit for chronic pain, including chronic musculoskeletal pain. A recent RCT showed that semaglutide improved knee pain associated with chronic knee osteoarthritis and obesity,^19^ and tirzepatide has been shown to produce statistically and clinically important improvement in bodily pain.^20^ While decreased mechanical stress, reduced adiposity (e.g., leptin levels), and anti-inflammatory mechanisms have all been implicated in pain amelioration pathways,^9,21^ such hypotheses have yet to be tested directly. Finally, despite encouraging findings related to GLP-1 RAs and pain outcomes, emerging evidence suggests that the effect of such treatments on chronic musculoskeletal pain could be bidirectional.^22^ For example, GLP-1 RAs can also lead to loss of lean muscle mass,^23^ which has been associated with increases in cLBP.^24^

In this pilot study, we investigated whether initiation of GLP-1 RAs is associated with improvements in pain severity, functional disability, quality of life, and physical performance among patients with obesity and cLBP. We hypothesized that patients would report reduced pain severity after 12 months of taking the medication. In addition, we aimed to explore potential mechanistic pathways by assessing changes in inflammatory biomarkers, adipokines, experimental pain sensitivity, and movement-evoked pain.

## Methods

### Design

This was a prospective cohort study of patients with obesity and cLBP taking GLP-1 RAs. All participants completed questionnaires at baseline, 3, 6, 9, and 12 months. At baseline and 6 months, those participating with in-person testing also underwent a blood draw, experimental pain sensitivity testing (quantitative sensory testing), and physical function testing. This study was approved by the Institutional Review Board at Washington University in St. Louis. All participants provided informed consent prior to study participation.

### Enrollment

Patients were eligible for inclusion if they were prescribed a GLP-1 RA (semaglutide or liraglutide) or a dual GLP-1/GIP agonist (tirzepatide) for treatment of obesity, were between 18 and 79 years old, and had a body mass index (BMI) between 35 and 50 kg/m^2^. They were also required to have cLBP, defined as pain which has persisted at least 3 months and had resulted in pain on at least half the days in the past 6 months,^25^ as well as a self-reported average low back pain intensity of ≥ 3/10 over the previous week. The upper BMI limit was chosen to increase the chance that a BMI decrease would lead to meaningful improvements in physical function.

Patients were excluded if they were being prescribed a GLP-1 RA primarily for glycemic control, had previous GLP-1 RA use, lumbar spine surgery, bariatric surgery, active malignancy, significant neurological disease, other chronic pain that the participant believed to be more severe than the cLBP, or back pain directly attributable to diagnosed structural pathology such as trauma, malignancy, infection, or ankylosing spondylitis. Participants were not eligible for in-person testing (quantitative sensory testing and blood draw) if they had uncontrolled hypertension (i.e. SBP/DBP of > 150/95), cardiovascular disease, or peripheral arterial disease. This exclusion was in place primarily for safety reasons as the cold pressor task represents a cardiovascular challenge, and additionally because hypertension can affect pain perception.^26^ This group of individuals was still eligible for participation in questionnaires.

Participants were recruited by contacting patients prescribed eligible medication at outpatient clinics affiliated with our hospital system over the previous week, or by self-referral. Recruitment lasted from July 2024 to January 2025, with a recruitment goal of at least 25 participants for this pilot study. After screening and informed consent, participants were scheduled for their first in-person visit either before or within two weeks after starting a GLP-1 RA. If participants were unable or unwilling to complete in-person testing, they were given the option of completing questionnaires only.

### Questionnaires

Participants completed online questionnaires at 3-month intervals for 12 months. Prior to study initiation, we initially assigned the Numerical Rating Scale (NRS) for back pain as the primary outcome. However, during the study’s conduct and prior to completing the final analysis, we chose to treat the Brief Pain Inventory-Short Form (BPI-SF) pain severity score as the primary outcome in this and future studies given its more comprehensive characterization of pain severity.^27^ The BPI-SF pain interference score and NRS pain scales for back, leg, neck, and arm pain were used as secondary outcomes related to pain severity across musculoskeletal regions. The Oswestry Disability Index (ODI) was used to assess back pain-related disability.^28^ To measure health-related quality of life, participants completed the 12-Item Short Form Health Survey v2 (SF-12), which yields a Physical Component Summary (PCS) and Mental Component Summary (MCS). Finally, participants self-reported their current weight and adherence to their GLP-1 RA medication.

### Inflammatory Biomarkers and Adipokines

At in-person visits at baseline and 6 months, participants underwent a blood draw. The inflammatory biomarkers high sensitivity C-reactive protein (hsCRP), tumor necrosis factor-alpha (TNF-α), interleukin-6 (IL-6), and interleukin-10 (IL-10) in serum were quantified. The hsCRP assay was performed on a Roche (Basel, Switzerland) cobas c501 system. TNF-α, IL-6 and IL-10 were measured using the SMCxPro instrument using commercially available kits from MilliporeSigma (Burlington, MA, USA). Samples also underwent analysis of the adipokines adiponectin and leptin using radioimmunoassay kits from Millipore (Burlington, MA, USA). Finally, hemoglobin A1c was measured in whole blood using the Roche (Basel, Switzerland) Tina Quant Gen3 assay on a cobas c501 system.

### Quantitative Sensory Testing

Quantitative sensory testing included both a mechanical temporal summation of pain (TSP) protocol and a conditioned pain modulation (CPM) protocol, both of which are well-established in the literature.^29–31^ Increased TSP indicates heightened pain facilitation, while increased CPM indicates increased pain inhibition.^31^

For TSP, punctate mechanical pain was assessed at the lumbar region (e.g., over erector spinae muscles) and the dorsal aspect of the non-dominant hand using a weighted (512 mN) pinprick stimulator (MRC Systems, Heidelberg, Germany). Participants provided a pain rating (0 = no pain, 100 = most intense pain imaginable) following a single contact of the punctate probe, after which they provided another pain rating following a series of 10 contacts at a rate of one contact per second. TSP is defined as the difference in pain ratings for the single versus multiple contacts.^29^

For CPM, the test stimulus was pressure pain applied to the lumbar region and forearm using an algometer (Medoc, Ramat Yishai, Israel). First, the amount of pressure required to elicit pain at baseline was assessed. Then, the participant was exposed to the conditioning stimulus, a cold pressor task in which the participant placed their hand in a 12°C water bath for as long as tolerable, up to 60 seconds. The amount of pressure required to elicit pain was then re-assessed following the conditioning stimulus. CPM was calculated as the percent change in pressure threshold following the conditioning stimulus compared to baseline.^32^

### Physical Function and Movement Evoked Pain Testing

Functional performance was assessed during in-person sessions using the Short Physical Performance Battery (SPPB), which consists of 1) standing balance, 2) 4-meter walking speed, and 3) chair rise tasks.^33^ After each of the 3 tasks, participants were asked to rate their pain while performing the task on a scale of 0 to 100 as a measure of movement-evoked pain.

### Analyses

Analyses were performed in R version 4.2.3 (R Foundation, Vienna, Austria). Results are reported throughout as median [Q1, Q3]. The Wilcoxon signed-rank test was used as a non-parametric test of significance for outcomes with only two timepoints. Linear mixed models with the fixed effect of treatment duration (month) were used to determine significance across multiple time points, with profile likelihood confidence intervals reported. To account for within-subject correlation and individual variability, random intercepts and slopes were used for each subject in the models. Missing data and loss to follow-up was handled implicitly through these models under the missing-at-random assumption. Boxplots display the median, first, and third quartiles, with whiskers representing the smallest/largest values within 1.5 times the interquartile range from the adjacent quartile, and dots displaying outliers.

To analyze correlations among outcomes, Spearman correlations were calculated based on the change in values from baseline, i.e., ΔBMI = BMI(6 months) – BMI(0 months). This analysis was conducted both for the outcomes available at 6 months, including quantitative sensory testing and blood biomarker testing, as well as for the more limited set of questionnaire outcomes available at 12 months.

## Results

### Participants

Out of 421 potentially eligible patients contacted, 35 started a GLP-1 RA and began participation in the study. Of these, 24 underwent in-person testing, and the remaining 11 were included for questionnaires only (Figure 1). At 6 months, questionnaire follow-up was completed for all participants, and in-person follow up was completed for 22 of 24. At 12 months, questionnaire follow-up was completed for 33 of 35 participants.

**Figure 1:**
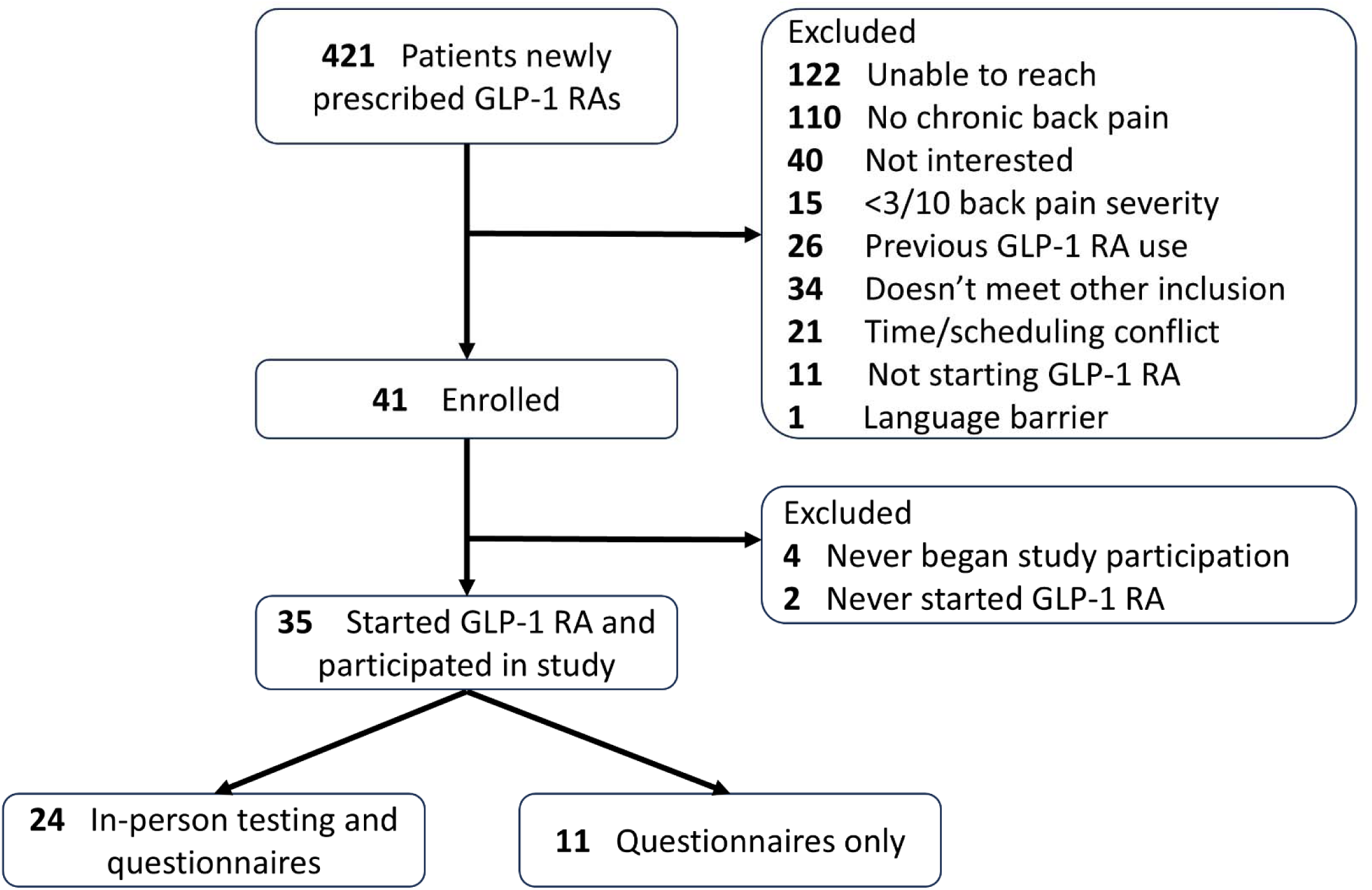
Enrollment flow chart.

Baseline participant characteristics are shown (Table 1). The median [Q1, Q3] baseline BMI was 39.9 [38.3, 43.6] kg/m^2^, and participants had an average NRS back pain of 6.0 [5.0, 7.0] over the prior week. Notably, the cohort was predominantly composed of women (86%). This imbalance was in part due to women making up a large proportion (69%) of the potentially eligible patients prescribed GLP-1 RAs. Twenty-four participants took tirzepatide, while 11 took semaglutide. No participants were prescribed liraglutide.

**Table 1:**
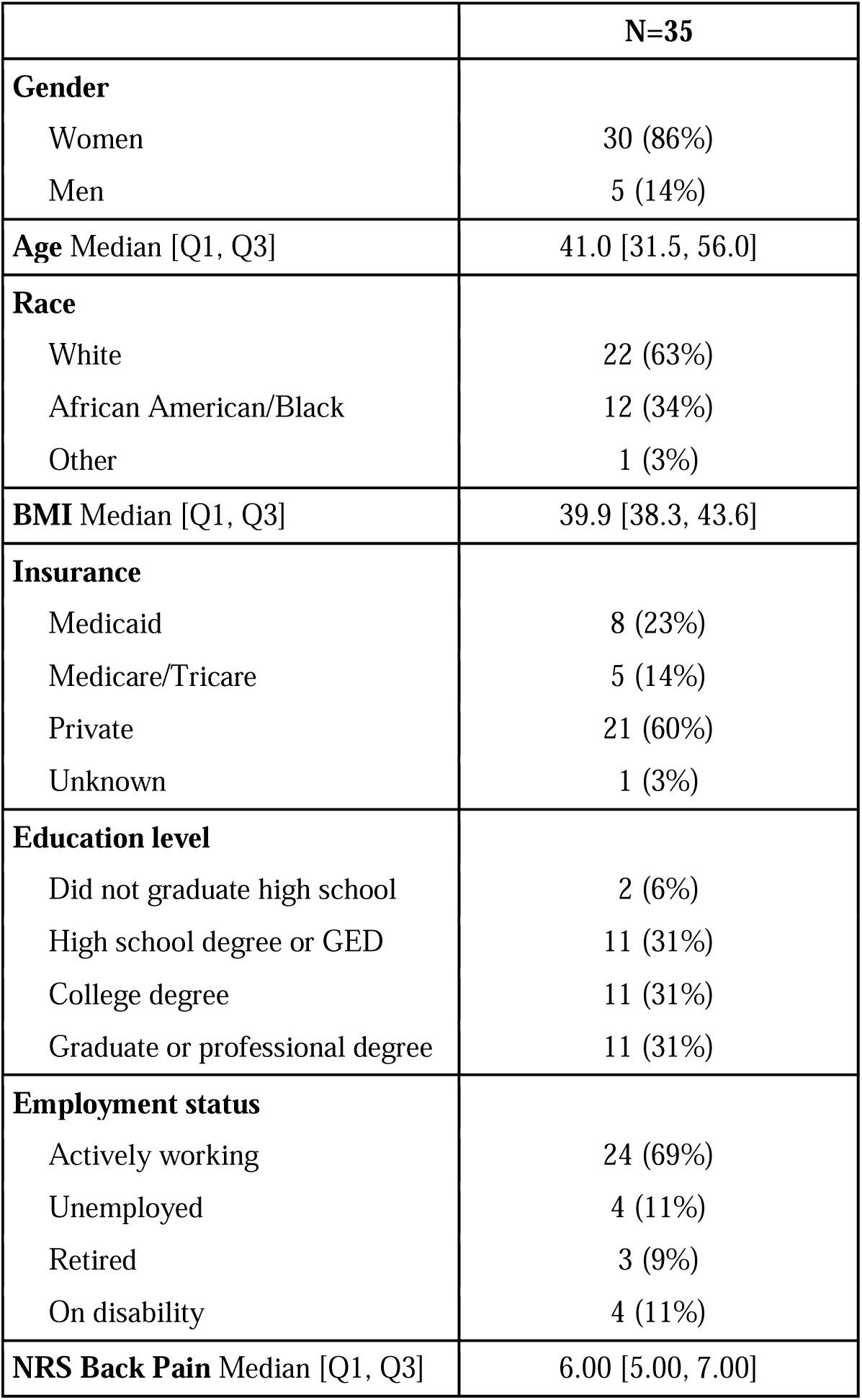
Baseline participant characteristics.

|  | **N=35** |
| --- | --- |
| **Gender** |  |
| Women | 30 (86%) |
| Men | 5 (14%) |
| **Age** Median [Q1, Q3] | 41.0 [31.5, 56.0] |
| **Race** |  |
| White | 22 (63%) |
| African American/Black | 12 (34%) |
| Other | 1 (3%) |
| **BMI** Median [Q1, Q3] | 39.9 [38.3, 43.6] |
| **Insurance** |  |
| Medicaid | 8 (23%) |
| Medicare/Tricare | 5 (14%) |
| Private | 21 (60%) |
| Unknown | 1 (3%) |
| **Education level** |  |
| Did not graduate high school | 2 (6%) |
| High school degree or GED | 11 (31%) |
| College degree | 11 (31%) |
| Graduate or professional degree | 11 (31%) |
| **Employment status** |  |
| Actively working | 24 (69%) |
| Unemployed | 4 (11%) |
| Retired | 3 (9%) |
| On disability | 4 (11%) |
| **NRS Back Pain** Median [Q1, Q3] | 6.00 [5.00, 7.00] |

### Questionnaires

From baseline to 12 months, BMI decreased by 12.5% from 39.9 [38.3, 43.6] kg/m^2^ to 34.9 [32.9, 39.8] kg/m^2^ (95% CI [-6.6, -4.2], p<.001) (Figure 2a). Over the same period, the primary outcome, BPI-SF pain severity, significantly decreased from 4.8 [2.9, 5.8] to 2.0 [1.0, 5.5] (95% CI [-2.1, -0.8], p<.001) (Figure 2b). Significant improvements were also observed for BPI-SF pain interference (95% CI [-2.2, -1.2]), ODI (95% CI [-10.5, - 4.4]), and SF-12 PCS (95% CI [4.3, 9.2]) (Figure 2c-e). Improvement in SF-12 MCS did not reach statistical significance (95% CI [0, 7.6]) (Figure 2f). NRS Back Pain significantly improved from baseline (95% CI [-3.3, -1.7]) (Figure 3a), as did NRS Leg Pain (95% CI [-2.5, -0.9]) (Figure 3b). NRS pain scores for neck and arm pain were less severe at baseline and did not reach significance (Figure 3c-d).

**Figure 2:**
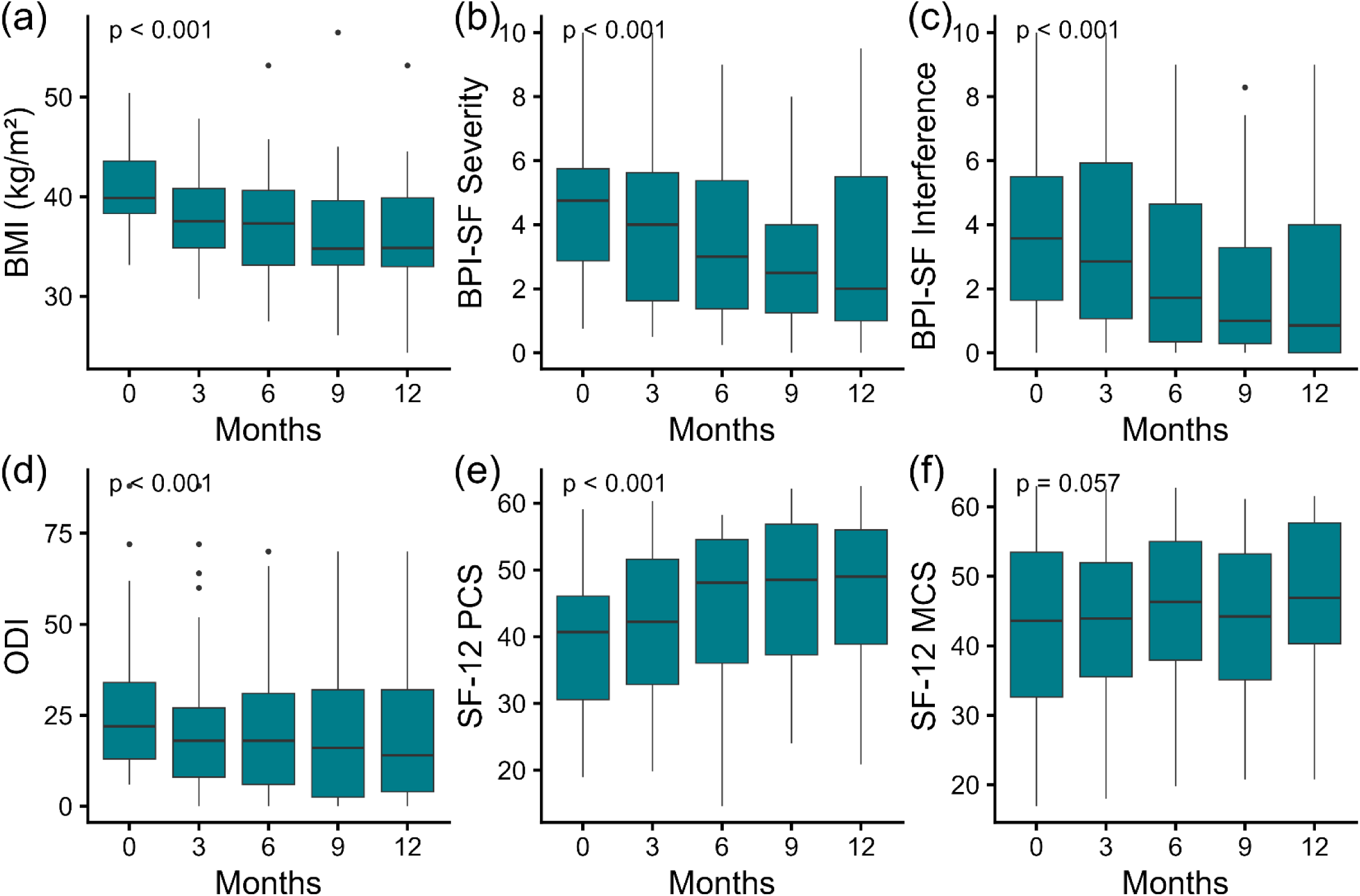
(a) Body mass index, (b) BPI-SF pain severity, (c) BPI-SF pain interference, (d) ODI, (e) SF-12 PCS, and (f) SF-12 MCS. The p-value corresponds to the fixed effect of treatment duration on the outcome in the linear mixed model.

**Figure 3:**
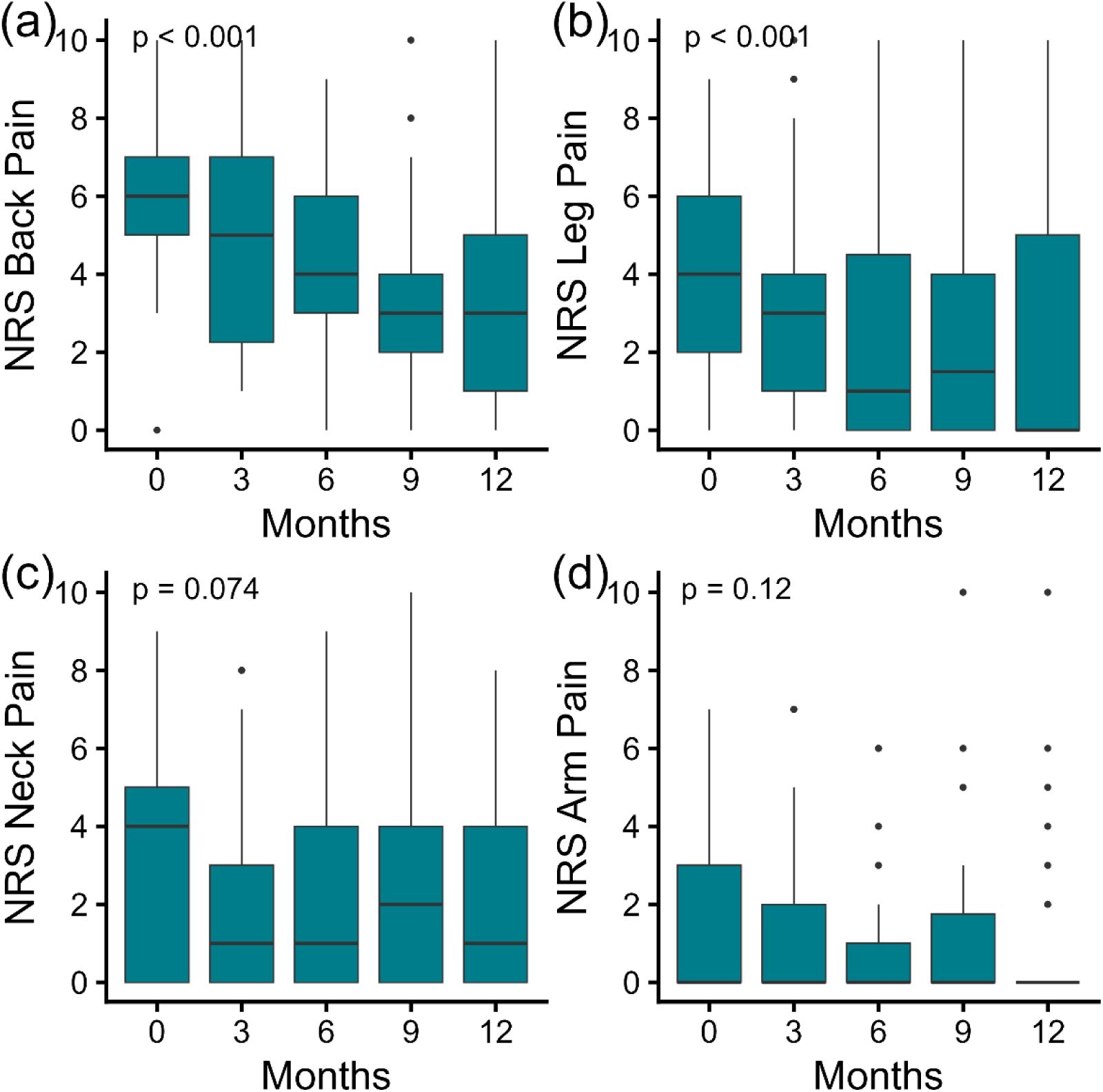
NRS pain scores for (a) back, (b) leg, (c) neck, and (d) arm.

Based on self-reported adherence, 20 participants never missed a dose, 9 rarely missed a dose, 3 sometimes missed a dose, and 3 rarely took the medication. BMI change at 12 months was clearly linked to adherence; the median BMI change at 1 year was -15% for those who never missed, -13% for those who rarely missed, -7% for those who sometimes missed, and +1% for those who rarely took the medication. Nearly two-thirds of participants (23/35) reported experiencing some side effects, most commonly nausea/vomiting (16), constipation (10), heartburn (5), diarrhea (4), and fatigue (4). By the end of 12 months, a total of seven participants had stopped taking the medication, primarily citing side effects and cost as reasons for discontinuation.

### Blood Testing

From baseline to 6 months, hemoglobin A1C significantly decreased from 5.5% [5.2%, 5.6%] to 5.2% [4.9%, 5.4%] (95% CI [-0.40, -0.20], p<.001) (Figure 4a). Over this period, the adipokines adiponectin and leptin both changed as anticipated with weight loss.^34^ Adiponectin increased from 6.2 [5.5, 8.3] µg/mL to 7.3 [5.5, 8.8] µg/mL (Figure 4b), though this did not reach significance (95% CI [-0.15, 1.22], p=.14). Leptin significantly decreased from 132 [88, 196] ng/mL to 71 [40, 140] ng/mL (95% CI [-63, -20], p<.01) (Figure 4c).

**Figure 4:**
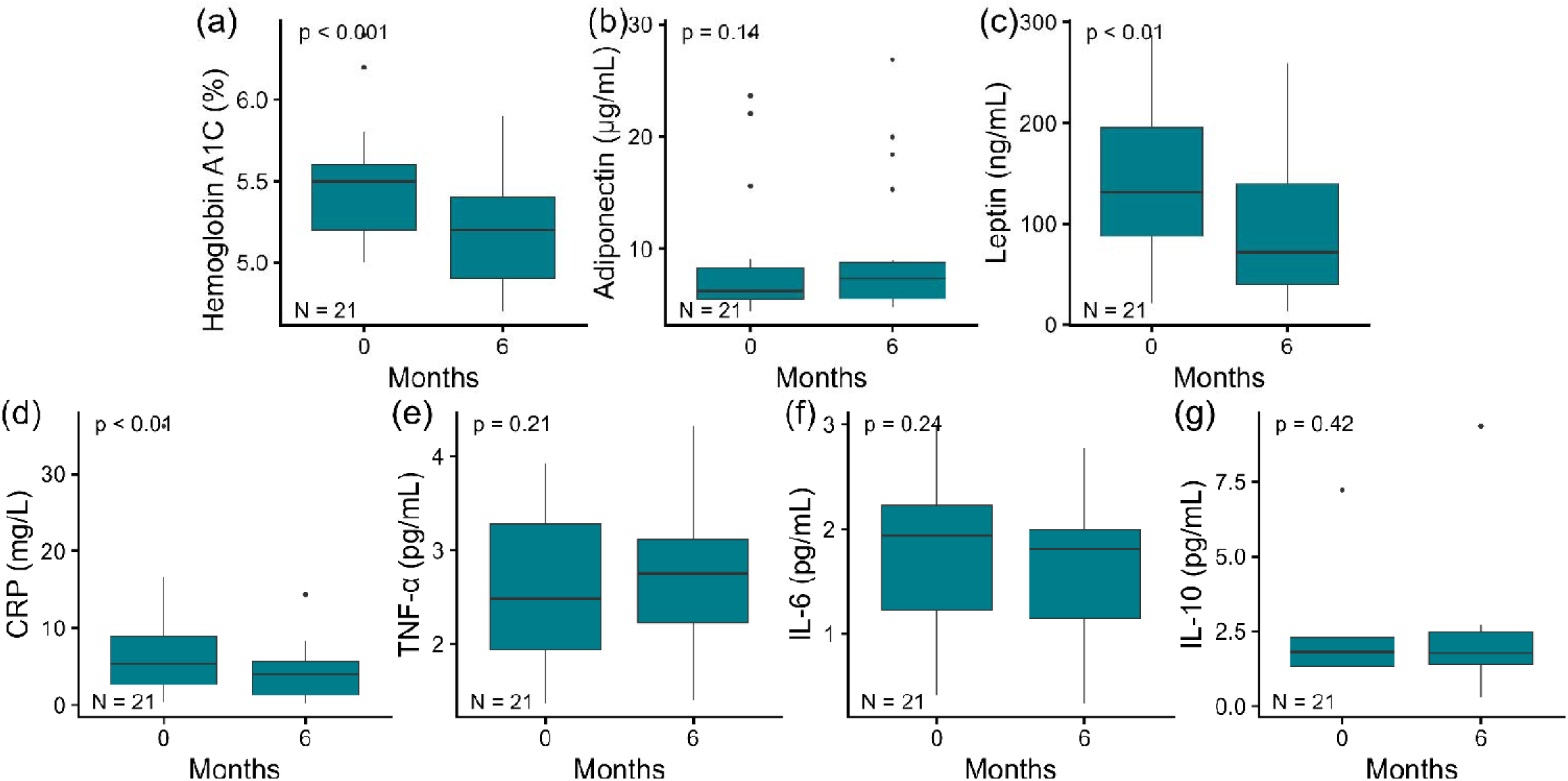
Blood test results at baseline and 6 months for (a) hemoglobin A1C, (b) adiponectin, (c) leptin, (d) C-reactive protein, (e) TNF-α, (f) IL-6, and (g) IL-10. P-values are calculated using the paired Wilcoxon signed-rank test.

There was a significant decrease observed in hsCRP, which fell from 5.4 [2.6, 8.9] mg/L to 4.0 [1.3, 5.7] mg/L (95% CI [-3.8, -0.6], p<.01) (Figure 4d). There were no significant changes in TNF-α, IL-6, or IL-10 (Figure 4e-g).

### Quantitative Sensory Testing and Physical Performance Testing

The median TSP value increased at both the hand and lower back from 0 to 6 months, but this was not statistically significant (Figure 5a). An increase in TSP would be consistent with increased pain facilitation.

**Figure 5:**
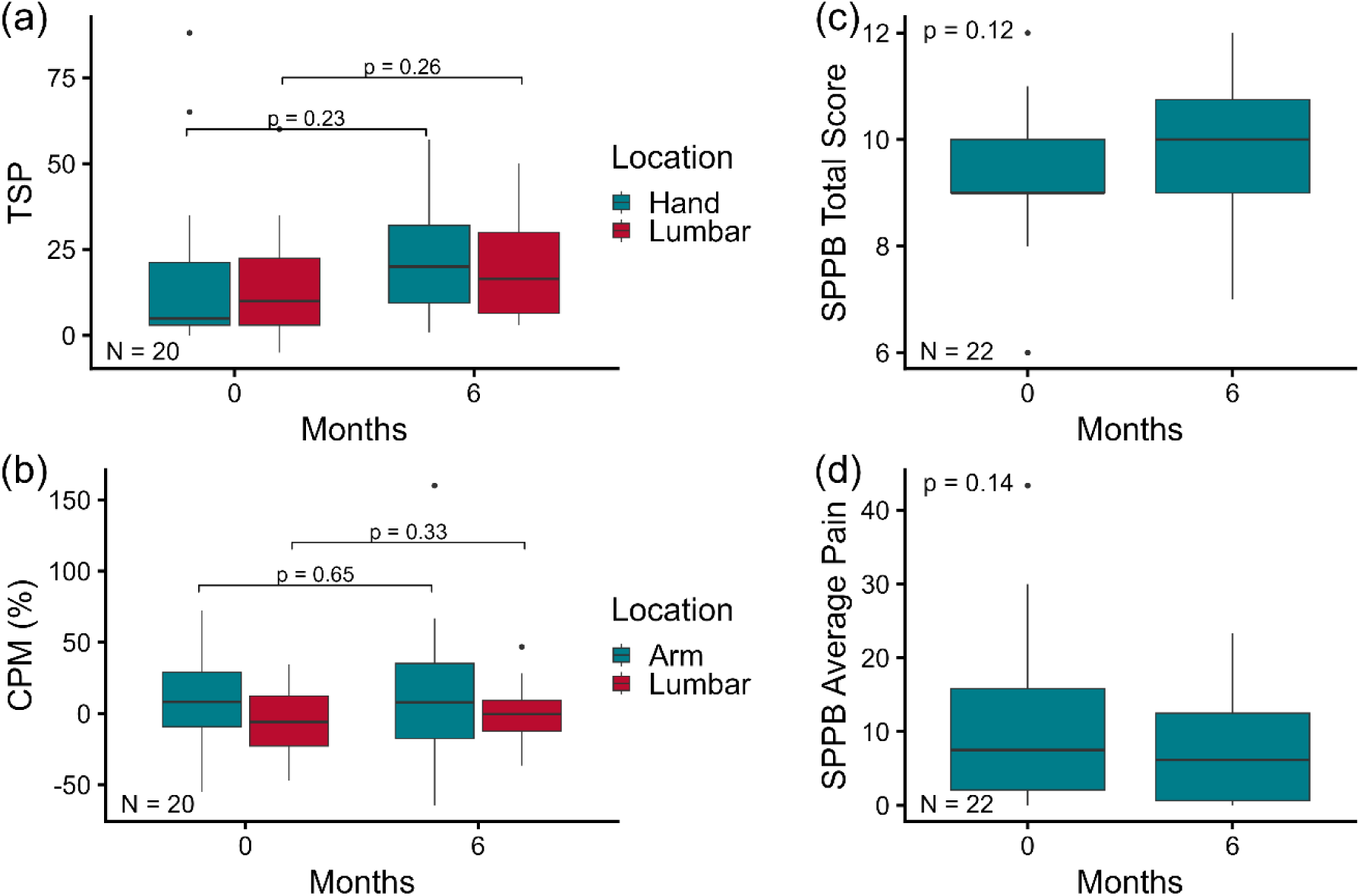
Quantitative sensory testing and physical performance testing results. (a) Temporal summation of pain (TSP) for hand and lumbar locations. (b) Conditioned pain modulation (CPM) for arm and lumbar locations. (c) Short physical performance battery (SPPB) total score. (d) Average pain level while performing the SPPB.

For CPM (Figure 5b), there was no change in pain inhibition at the arm location (Baseline: 8.2%, 6 months: 7.7%, p=.65). Here, a positive value for CPM indicates an increased pain threshold when the conditioning stimulus was applied, as expected. However, for the lumbar location, the pain threshold decreased with the addition of the conditioning stimulus at both timepoints, resulting in negative values. There was a trend toward a more positive CPM at the lumbar location from 0 to 6 months (Baseline: -6.0%, 6 months: -0.4%, p=.33), which would indicate increased pain inhibition, but this was not significant.

For physical performance testing with the SPPB, the median score increased from 9 at baseline to 10 at 6 months, but this did not reach significance (95% CI [0.0, 1.5], p=.12) (Figure 5c). Similarly, the participants’ average self-reported movement-evoked pain across the 3 tasks decreased from a median of 7.5 to 6.2 but did not reach significance (95% CI [-10.5, 1.7], p=.14) (Figure 5d).

### Correlations

At both 6 and 12 months, the changes among pain scores (ΔBPI-SF severity, ΔBPI-SF interference, ΔNRS Back) were moderately correlated, with r_s_ ranging from .44 to .67 (Figure 6). At 6 months, ΔCRP was moderately correlated with both ΔBPI-SF severity (r_s_ = .49) and ΔBMI (r_s_ = .47). ΔLeptin and ΔBMI were also significantly correlated (r_s_ = .68). Interestingly, ΔBMI at 12 months was not significantly correlated with ΔBPI-SF severity (r_s_ = .27), though it was significantly correlated with ΔODI (r_s_ = .42).

**Figure 6:**
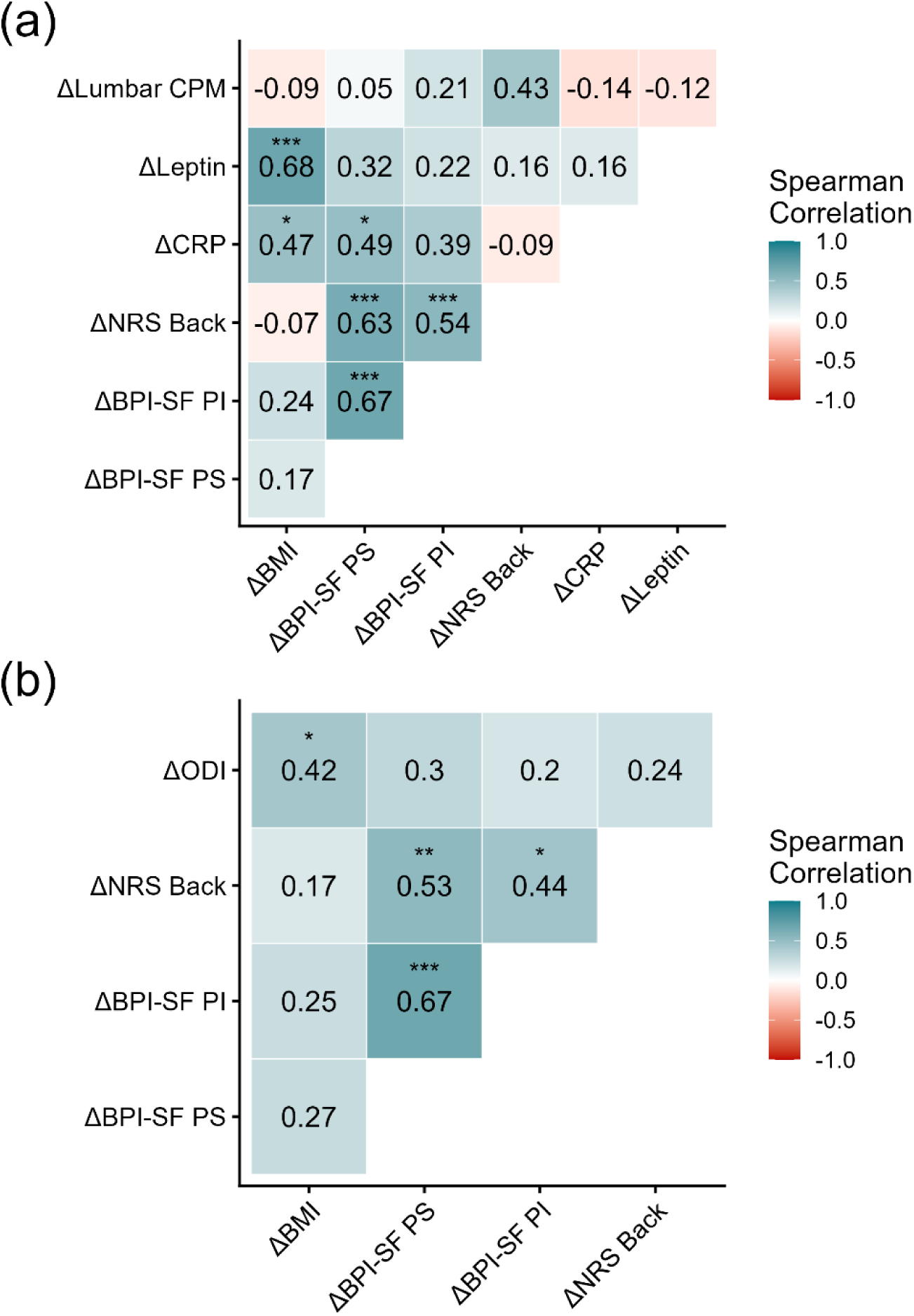
Spearman correlations among the change (Δ) in outcomes from baseline to (a) 6 months and (b) 12 months. *p<0.05; **p<0.01, ***p<0.001. PS=pain severity. PI=pain interference.

## Discussion

In this prospective pilot study, patients with obesity and cLBP starting GLP-1 RAs experienced clinically meaningful improvements in pain severity, functional disability, and health-related quality of life over 12 months. These findings further extend the potential therapeutic relevance of GLP-1 RAs beyond metabolic disease and suggest that these medications may have a role in pain management for patients with obesity. In addition to reductions in pain, the cohort experienced decreased BMI, hemoglobin A1c, leptin, and CRP, pointing to both metabolic and inflammatory pathways as plausible contributors.

The clinical improvements observed in this study are compelling and suggestive of a meaningful therapeutic benefit. The median reduction in NRS Back Pain severity was 3 points over 12 months, which exceeds an accepted threshold for minimum clinically important difference in chronic low back pain.^35^ Furthermore, reductions in disability and improvements in quality of life were consistent across multiple validated instruments. Compared to other nonsurgical interventions, the magnitude of benefit appears similar or greater. For example, exercise therapy results in small to moderate effects, with an average improvement of 1.5 points.^36^ Cognitive behavioral therapy and related interventions also produce only small improvements, with significant variability across trials.^37^ Bariatric surgery cohorts have also demonstrated meaningful reductions in back pain severity, with a meta-analysis finding a mean effect size of 3 points on the NRS or Visual Analog Scale.^38^ While surgery is generally associated with greater weight loss compared to GLP-1 RAs and does not require adherence to medication,^39^ it comes at the cost of an invasive procedure. While these findings suggest that GLP-1 RAs may offer clinically meaningful relief of cLBP, it is important to acknowledge limitations of this therapy, including high cost, potential need for long-term use, and gastrointestinal side effects that can affect tolerability.^40^ Furthermore, long-term safety and durability of benefits remain uncertain.

While the questionnaire results provide compelling evidence that back pain severity can significantly improve with GLP-1 RAs, other aspects require additional investigation. Experimental pain sensitivity testing did not show clear evidence of either decreased pain facilitation or increased pain inhibition, contrary to some research suggesting that GLP-1 RAs may suppress pain hypersensitivity.^41^ Both physical performance testing and measures of movement-evoked pain did appear to improve, though this did not reach significance in this small sample size. While CRP decreased significantly from baseline to 6 months, TNF-α, IL-6, and IL-10 were not significantly changed. Decreases in CRP have been more commonly associated with weight loss interventions in the literature,^18,42^ despite some studies demonstrating changes in TNF-α, IL-6, or IL-10.^43,44^ Interestingly, there was not a significant correlation between the magnitudes of change in BMI and change in pain scores among individuals, despite significant reductions in both across the cohort as a whole. Inflammation, quantified by CRP, was significantly correlated with BPI-SF pain severity. Future studies should further examine the mechanistic pathways by which GLP-1 RAs affect cLBP outcomes.

Multiple studies have investigated the effects of weight loss and healthy lifestyle interventions on cLBP, but recent systematic reviews have identified only low-quality evidence suggesting that these may have an impact on back pain and disability.^14,45^ As a consequence of this lack of evidence, the World Health Organization guidelines on cLBP management do not currently recommend weight loss interventions as a treatment strategy.^46^ Single-arm studies following patients receiving bariatric surgery have observed reduced back pain as well as increased disc space height,^12,47^ but RCTs have not been conducted because of the invasiveness and associated risk of the procedure. A single-arm pilot study for a medically supervised weight loss program by Roffey et al. found significant improvements in pain and disability compared to baseline, but noted issues with program adherence and maintenance of weight loss.^13^ An RCT of a healthy lifestyle intervention by Williams et al. found no impact on pain intensity, but the intervention also failed to produce weight loss relative to control.^48^

A more recent RCT found evidence of modest improvement (-1.3 point-difference; 24 point scale) in back-pain related disability with a multidimensional lifestyle intervention that included nutrition and weight management advice.^49^ However, there was no difference in pain severity associated with the intervention, and most notably, effects were no different between patients who had elevated versus normal BMI. Weight loss was also minimal (difference of -1.6 kg in the treatment group), emphasizing a key shortcoming of the existing literature: except for bariatric surgery, most lifestyle and pharmacologic interventions given to patients with cLBP and obesity have, until recently, not led to meaningful and reliable weight loss.

The recent development of GLP-1 RAs as highly effective obesity pharmacotherapies has for the first time allowed for reliable nonsurgical weight loss. To our knowledge, this is the first prospective study to investigate the association between GLP-1 RA use and back pain. In a retrospective study of the US Department of Veterans Affairs database, Xie et al. found no significant effect of these medications on the subsequent incidence of a low back pain diagnosis.^22^ Despite the strengths of that study, a significant limitation is that diagnosis codes in the electronic health record have limited sensitivity for cLBP,^50^ and these diagnoses do not capture severity or changes over time. Instead, our prospective study utilized validated pain questionnaires administered at multiple timepoints.

### Limitations

While this study addresses a novel and timely research question, it has multiple limitations. This was a single-arm, observational study, and it is therefore impossible to establish a definite causal relationship. This study was designed as a proof-of-concept investigation and not a clinical trial. The absence of a control group is a fundamental limitation; without a comparator arm, it is not possible to determine whether the observed improvements reflect the effect of GLP-1 RAs or alternative explanations such as natural disease history, regression to the mean, or the placebo effect. While included participants were required to have low back pain that was chronic in nature, it is possible their pain severity spontaneously improved over the study duration. Readers should therefore interpret these preliminary findings with appropriate caution.

This was also a pilot study with only 35 participants, limiting statistical power. The study participants were predominantly women, which reflected prescribing patterns but limits the generalizability of these findings. Finally, this study did not include spine imaging, which could have yielded useful information on the relative efficacy for participants who had identifiable pathology of the lumbar spine (e.g., disc herniation or spondylolisthesis) compared to those with nonspecific axial back pain.

## Conclusion

This pilot study provides early evidence that GLP-1 RAs could be a promising new therapy for the management of cLBP in patients with obesity. While it is expected that addressing a risk factor for back pain should improve the severity of that pain, this assumption has previously been difficult to experimentally prove due to the difficulty of achieving weight loss without an invasive procedure like bariatric surgery. By helping patients achieve reliable, nonsurgical weight loss, GLP-1 RAs could help many struggling with both obesity and cLBP. Building on these proof-of-concept findings, we are currently planning a larger RCT to establish causality and further investigate the mechanisms underlying these clinical changes.

## Acknowledgements

Blood samples were run by the Core Laboratory for Clinical Studies at Washington University, with contributions from Dr. Jennifer Powers Carson and Meghan Horvath.

## AI statement

During the preparation of this work the authors used Claude by Anthropic to improve clarity and conciseness of the manuscript. After using this tool, the authors reviewed and edited the content as needed and take full responsibility for the content of the published article.

## Funding disclosure

This work was supported by The Foundation for Barnes-Jewish Hospital. Additional financial support for laboratory testing was received from the NIH Shared Instrumentation Grant #S10OD027006 and the Washington University Diabetes Research Center NIH P30 Grant #DK020579.

## Conflicts of interest

Dr. Greenberg has received research support paid to the institution from Medtronic and has consulted for Medtronic and Kuros Biosciences. Dr. Goodin is the current president of the United States Association for the Study of Pain. The remaining authors have no conflicts to disclose.

## Data availability

The data that supports the findings of this study are available from the corresponding author upon reasonable request.

## Notes

### Author Declarations

IRB of Washington University School of Medicine in St. Louis gave ethical approval for this work.

## References

1. GBD 2021 Low Back Pain Collaborators. Global, regional, and national burden of low back pain, 1990-2020, its attributable risk factors, and projections to 2050: a systematic analysis of the Global Burden of Disease Study 2021. Lancet Rheumatol. 2023;5(6):e316-e329. doi:10.1016/S2665-9913(23)00098-X

2. Dieleman JL, Cao J, Chapin A, et al. US Health Care Spending by Payer and Health Condition, 1996-2016. JAMA. 2020;323(9):863-884. doi:10.1001/jama.2020.0734

3. Ricci JA, Stewart WF, Chee E, Leotta C, Foley K, Hochberg MC. Back Pain Exacerbations and Lost Productive Time Costs in United States Workers. Spine. 2006;31(26):3052. doi:10.1097/01.brs.0000249521.61813.aa

4. Jenkins HJ, Corrêa L, Brown BT, et al. Long-term effectiveness of non-surgical interventions for chronic low back pain: a systematic review and meta-analysis. Lancet Rheumatol. 2025;7(9):e607–e617. doi:10.1016/S2665-9913(25)00064-5

5. Shiri R, Karppinen J, Leino-Arjas P, Solovieva S, Viikari-Juntura E. The Association Between Obesity and Low Back Pain: A Meta-Analysis. Am J Epidemiol. 2010;171(2):135–154. doi:10.1093/aje/kwp356

6. Chen N, Fong DYT, Wong JYH. Health and Economic Outcomes Associated With Musculoskeletal Disorders Attributable to High Body Mass Index in 192 Countries and Territories in 2019. JAMA Netw Open. 2023;6(1):e2250674. doi:10.1001/jamanetworkopen.2022.50674

7. Xu X, Li X, Wu W. Association Between Overweight or Obesity and Lumbar Disk Diseases: A Meta-Analysis. Clin Spine Surg. 2015;28(10):370. doi:10.1097/BSD.0000000000000235

8. Segar AH, Baroncini A, Urban JPG, Fairbank J, Judge A, McCall I. Obesity increases the odds of intervertebral disc herniation and spinal stenosis; an MRI study of 1634 low back pain patients. Eur Spine J. 2024;33(3):915–923. doi:10.1007/s00586-024-08154-4

9. Binvignat M, Sellam J, Berenbaum F, Felson DT. The role of obesity and adipose tissue dysfunction in osteoarthritis pain. Nat Rev Rheumatol. 2024;20(9):565–584. doi:10.1038/s41584-024-01143-3

10. Francisco V, Pino J, González-Gay MÁ, et al. A new immunometabolic perspective of intervertebral disc degeneration. Nat Rev Rheumatol. 2022;18(1):47–60. doi:10.1038/s41584-021-00713-z

11. van den Berg R, Jongbloed EM, de Schepper EIT, Bierma-Zeinstra SMA, Koes BW, Luijsterburg PAJ. The association between pro-inflammatory biomarkers and nonspecific low back pain: a systematic review. Spine J. 2018;18(11):2140–2151. doi:10.1016/j.spinee.2018.06.349

12. Khoueir P, Black MH, Crookes PF, Kaufman HS, Katkhouda N, Wang MY. Prospective assessment of axial back pain symptoms before and after bariatric weight reduction surgery. Spine J. 2009;9(6):454–463. doi:10.1016/j.spinee.2009.02.003

13. Roffey DM, Ashdown LC, Dornan HD, et al. Pilot evaluation of a multidisciplinary, medically supervised, nonsurgical weight loss program on the severity of low back pain in obese adults. Spine J. 2011;11(3):197–204. doi:10.1016/j.spinee.2011.01.031

14. Chen LH, Weber K, Mehrabkhani S, Baskaran S, Abbass T, Macedo LG. The effectiveness of weight loss programs for low back pain: a systematic review. BMC Musculoskelet Disord. 2022;23(1):488. doi:10.1186/s12891-022-05391-w

15. Wilding JPH, Batterham RL, Calanna S, et al. Once-Weekly Semaglutide in Adults with Overweight or Obesity. N Engl J Med. 2021;384(11):989–1002. doi:10.1056/NEJMoa2032183

16. Jastreboff AM, Aronne LJ, Ahmad NN, et al. Tirzepatide Once Weekly for the Treatment of Obesity. N Engl J Med. 2022;387(3):205–216. doi:10.1056/NEJMoa2206038

17. Badve SV, Bilal A, Lee MMY, et al. Effects of GLP-1 receptor agonists on kidney and cardiovascular disease outcomes: a meta-analysis of randomised controlled trials. Lancet Diabetes Endocrinol. 2025;13(1):15–28. doi:10.1016/S2213-8587(24)00271-7

18. Verma S, Bhatta M, Davies M, et al. Effects of once-weekly semaglutide 2.4 mg on C-reactive protein in adults with overweight or obesity (STEP 1, 2, and 3): Exploratory analyses of three randomised, double-blind, placebo-controlled, phase 3 trials. EClinicalMedicine. 2023;55:101737. doi:10.1016/j.eclinm.2022.101737

19. Bliddal H, Bays H, Czernichow S, et al. Once-Weekly Semaglutide in Persons with Obesity and Knee Osteoarthritis. N Engl J Med. 2024;391(17):1573–1583. doi:10.1056/NEJMoa2403664

20. Hunter Gibble T, Cao D, Zhang XM, Xavier NA, Poon JL, Fitch A. Tirzepatide Was Associated with Improved Health-Related Quality of Life in Adults with Obesity or Overweight and Type 2 Diabetes: Results from the Phase 3 SURMOUNT-2 Trial. Diabetes Ther. 2025;16(5):977–991. doi:10.1007/s13300-025-01723-w

21. Felson DT. Glucagon-Like Peptide-1 Receptor Agonists and Osteoarthritis. N Engl J Med. 2024;391(17):1643–1644. doi:10.1056/NEJMe2409972

22. Xie Y, Choi T, Al-Aly Z. Mapping the effectiveness and risks of GLP-1 receptor agonists. Nat Med. Published online January 20, 2025:1–12. doi:10.1038/s41591-024-03412-w

23. Bikou A, Dermiki-Gkana F, Penteris M, Constantinides TK, Kontogiorgis C. A systematic review of the effect of semaglutide on lean mass: insights from clinical trials. Expert Opin Pharmacother. 2024;25(5):611–619. doi:10.1080/14656566.2024.2343092

24. Fortin M, Macedo LG. Multifidus and paraspinal muscle group cross-sectional areas of patients with low back pain and control patients: a systematic review with a focus on blinding. Phys Ther. 2013;93(7):873–888. doi:10.2522/ptj.20120457

25. Deyo RA, Dworkin SF, Amtmann D, et al. Report of the NIH Task Force on research standards for chronic low back pain. J Pain. 2014;15(6):569–585. doi:10.1016/j.jpain.2014.03.005

26. Sheps DS, Bragdon EE, Gray TF, Ballenger M, Usedom JE, Maixner W. Relation between systemic hypertension and pain perception. Am J Cardiol. 1992;70(16):F3–F5. doi:10.1016/0002-9149(92)90181-W

27. Tan G, Jensen MP, Thornby JI, Shanti BF. Validation of the Brief Pain Inventory for chronic nonmalignant pain. J Pain. 2004;5(2):133–137. doi:10.1016/j.jpain.2003.12.005

28. Fairbank JC, Pynsent PB. The Oswestry Disability Index. Spine. 2000;25(22):2940–2952; discussion 2952. doi:10.1097/00007632-200011150-00017

29. Goodin BR, Bulls HW, Herbert MS, et al. Temporal summation of pain as a prospective predictor of clinical pain severity in adults aged 45 years and older with knee osteoarthritis: ethnic differences. Psychosom Med. 2014;76(4):302–310. doi:10.1097/PSY.0000000000000058

30. Yarnitsky D, Bouhassira D, Drewes AM, et al. Recommendations on practice of conditioned pain modulation (CPM) testing. Eur J Pain. 2015;19(6):805–806. doi:10.1002/ejp.605

31. Horn-Hofmann C, Kunz M, Madden M, Schnabel EL, Lautenbacher S. Interactive effects of conditioned pain modulation and temporal summation of pain—the role of stimulus modality. PAIN. 2018;159(12):2641. doi:10.1097/j.pain.0000000000001376

32. Goodin BR, Anderson AJB, Freeman EL, Bulls HW, Robbins MT, Ness TJ. Intranasal oxytocin administration is associated with enhanced endogenous pain inhibition and reduced negative mood states. Clin J Pain. 2015;31(9):757–767. doi:10.1097/AJP.0000000000000166

33. Guralnik JM, Simonsick EM, Ferrucci L, et al. A short physical performance battery assessing lower extremity function: association with self-reported disability and prediction of mortality and nursing home admission. J Gerontol. 1994;49(2):M85–94. doi:10.1093/geronj/49.2.m85

34. Zhao S, Kusminski CM, Scherer PE. Adiponectin, Leptin and Cardiovascular Disorders. Circ Res. 2021;128(1):136-149. doi:10.1161/CIRCRESAHA.120.314458

35. Suzuki H, Aono S, Inoue S, et al. Clinically significant changes in pain along the Pain Intensity Numerical Rating Scale in patients with chronic low back pain. PLOS ONE. 2020;15(3):e0229228. doi:10.1371/journal.pone.0229228

36. Hayden JA, Ellis J, Ogilvie R, Malmivaara A, Tulder MW van. Exercise therapy for chronic low back pain. Cochrane Database Syst Rev. 2021;2021(9). doi:10.1002/14651858.cd009790.pub2

37. Richmond H, Hall AM, Copsey B, et al. The Effectiveness of Cognitive Behavioural Treatment for Non-Specific Low Back Pain: A Systematic Review and Meta-Analysis. PLOS ONE. 2015;10(8):e0134192. doi:10.1371/journal.pone.0134192

38. Stefanova I, Currie AC, Newton RC, et al. A Meta-analysis of the Impact of Bariatric Surgery on Back Pain. Obes Surg. 2020;30(8):3201–3207. doi:10.1007/s11695-020-04713-y

39. Barrett TS, Hafermann JO, Richards S, LeJeune K, Eid GM. Obesity Treatment With Bariatric Surgery vs GLP-1 Receptor Agonists. JAMA Surg. 2025;160(11):1232–1239. doi:10.1001/jamasurg.2025.3590

40. Thomsen RW, Mailhac A, Løhde JB, Pottegård A. Real-world evidence on the utilization, clinical and comparative effectiveness, and adverse effects of newer GLP-1RA-based weight-loss therapies. Diabetes Obes Metab. 2025;27(S2):66–88. doi:10.1111/dom.16364

41. He Y, Xu B, Zhang M, et al. Advances in GLP-1 receptor agonists for pain treatment and their future potential. J Headache Pain. 2025;26(1):46. doi:10.1186/s10194-025-01979-4

42. Pardina E, Ferrer R, Baena-Fustegueras JA, et al. Only C-reactive protein, but not TNF-α or IL6, reflects the improvement in inflammation after bariatric surgery. Obes Surg. 2012;22(1):131–139. doi:10.1007/s11695-011-0546-3

43. Bulmer C, Avenell A. The effect of dietary weight-loss interventions on the inflammatory markers interleukin-6 and TNF-alpha in adults with obesity: A systematic review and meta-analysis of randomized controlled clinical trials. Obes Rev Off J Int Assoc Study Obes. 2025;26(7):e13910. doi:10.1111/obr.13910

44. Jung SH, Park HS, Kim KS, et al. Effect of weight loss on some serum cytokines in human obesity: increase in IL-10 after weight loss. J Nutr Biochem. 2008;19(6):371–375. doi:10.1016/j.jnutbio.2007.05.007

45. Huijbers JCJ, Coenen P, Burchell GLB, et al. The (cost-)effectiveness of combined lifestyle interventions for people with persistent low-back pain who are overweight or obese: A systematic review. Musculoskelet Sci Pract. 2023;65:102770. doi:10.1016/j.msksp.2023.102770

46. WHO Guideline for Non-Surgical Management of Chronic Primary Low Back Pain in Adults in Primary and Community Care Settings. World Health Organization; 2023. Accessed November 26, 2025. http://www.ncbi.nlm.nih.gov/books/NBK599212/

47. Lidar Z, Behrbalk E, Regev GJ, et al. Intervertebral disc height changes after weight reduction in morbidly obese patients and its effect on quality of life and radicular and low back pain. Spine. 2012;37(23):1947–1952. doi:10.1097/BRS.0b013e31825fab16

48. Williams A, Wiggers J, O’Brien KM, et al. Effectiveness of a healthy lifestyle intervention for chronic low back pain: a randomised controlled trial. Pain. 2018;159(6):1137–1146. doi:10.1097/j.pain.0000000000001198

49. Mudd E, Davidson SRE, Kamper SJ, et al. Healthy Lifestyle Care vs Guideline-Based Care for Low Back Pain: A Randomized Clinical Trial. JAMA Netw Open. 2025;8(1):e2453807. doi:10.1001/jamanetworkopen.2024.53807

50. Ly A, Sirois C, Dionne CE. Sensitivity and specificity of algorithms for the identification of nonspecific low back pain in medico-administrative databases. Pain. 2023;164(7):1600–1607. doi:10.1097/j.pain.0000000000002861

